# Associations of childcare type, age at start, and intensity with body mass index trajectories from 10 to 42 years of age in the 1970 British Cohort Study

**DOI:** 10.1101/19003772

**Authors:** Silvia Costa, David Bann, Sara E Benjamin-Neelon, Jean Adams, William Johnson

## Abstract

**Background:** Attending childcare is related to greater childhood obesity risk, but there are few long-term follow-up studies. We aimed to examine the associations of childcare type, duration, and intensity with BMI trajectories from ages 10-42 years.

**Methods:** The sample comprised 8234 individuals in the 1970 British Cohort Study, who had data on childcare attendance (no, yes), type (formal, informal), duration (4-5, 3-3.99, 0-2.99 years old when started), and intensity (1, 2, 3, 4-5 days/week) reported at age five years and 32563 BMI observations. Multilevel linear spline models were used to estimate the association of each exposure with the sample-average BMI trajectory, with covariate adjustment. A combined duration and intensity exposure was also examined.

**Results:** Childcare attendance and type were not strongly related to BMI trajectories. Among participants who attended childcare 1-2 days a week, those who started when 3-3.99 years old had a 0.197 (-0.004, 0.399) kg/m^2^ higher BMI at age 10 years than those who started when 4-5 years old, and those who started when 0-2.99 years old had a 0.289 (0.049, 0.529) kg/m^2^ higher BMI. A similar dose-response pattern for intensity was observed when holding duration constant. By age 42 years, individuals who started childcare at age 0-2.99 years and attended 3-5 days/week had a 1.356 kg/m^2^ (0.637, 2.075) higher BMI than individuals who started at age 4-5 years and attended 1-2 days/week.

**Conclusions:** Children who start childcare earlier and/ or attend more frequently may have greater long-term obesity risk.

## Introduction

Obesity is a major public health problem, accounting for four million deaths globally and 120 million disability-adjusted life-years in 2015.(1) In the United Kingdom (UK), approximately 25% of the adulthood population are currently obese, with 40% of men and 30% of women overweight.(2) Even more worrying is that more recent generations are developing obesity at increasingly younger ages.(3) Because obesity tracks from childhood to adulthood(4), this means that future rates of obesity in adulthood are projected to increase.(5)

Childcare settings (e.g., nurseries and playgroups) may offer an ideal opportunity for population-level interventions and targeted policy changes. Namely because healthy behaviours and obesogenic trajectories are established early in life(3,6-13) and a rising number of children attend out-of-home childcare, with many starting as early as the first year of life and spending much of their week days in these settings.(14-16) Two recent reviews have summarised the literature on the association between exposure to childcare, one between ages 0-2 years(17) and one between ages 0-5 years,(18) and obesity risk. Both these publications found that available evidence is limited to cross-sectional and short-term follow-up studies, with most studies either revealing no association or a harmful effect. There is some evidence that earlier age of starting childcare (indicating greater duration of exposure) and higher intensity of attendance are particularly associated with greater obesity. While certain aspects of childcare may be related to increased risk for obesity in childhood, we know almost nothing about how exposure to childcare might be associated with body mass index (BMI) into and across adulthood. Such knowledge would provide evidence that targeting childcare policies and practices might ultimately benefit adulthood health and wellbeing.

We aimed to examine the associations of childcare attendance, and type, age at start, and intensity among those children who did attend, with BMI trajectories from 10 to 42 years of age in the 1970 British Birth Cohort.

## Methods

### Study sample

The 1970 British Cohort Study is based on 17198 people born in one week in April 1970 in England, Scotland, and Wales.(19) Data collections have taken place at ages 0 (1970), 5 (1975), 10 (1980), 16 (1986), 26 (1996), 30 (2000), 34 (2004), and 42 (2012) years. The study has received ethical approval and obtained informed parental and/or participant consent for all data collection. At the most recent sweep, the 9841 individuals still participating in the study remained broadly representative of the national population of men and women of the same age.

Starting with the 14875 cohort members who were still participating at age 10 years (i.e., when BMI was first assessed), 4843 were excluded because of missing childcare data, 608 because they did not have a single BMI measurement, and 1190 because of missing data on potential confounders. The resulting sample comprised 8234 individuals, representing 55% of the eligible cohort (i.e., N = 14875).

### Data

#### Outcome

Weight and height were measured by community medical officers, health visitors, or school nurses at ages 10 and 16 years according to standard protocols. At the adulthood sweeps, weight and height were self-reported in questionnaires (age 26 years) or face-to-face interviews (ages 30, 34, and 42 years). BMI was computed as weight (kg) / height (m^2^). In total there were 32563 observations of BMI. Approximately 65% of the sample had four or more observations (out of a maximum of six) and approximately 72% of the sample had serial BMI measurements spanning more than 20 years (out of a maximum of 32 years).

#### Exposures

At age five years, mothers reported type of childcare setting, age when the participant started, and how many morning and afternoon sessions per week the participant attended. Because these data were collected only for the most recent and previous main childcare placements that lasted for three months or longer, our exposure variable is limited to the main childcare setting attended by the participant. The first exposure we computed was attendance: “no” if all placements were coded as does not attend or “yes” if any placement was coded as does attend. The second exposure was type: “formal” for nursery school, nursery class attached to a school, or nursery or “informal” for playgroup. The “informal” classification is restricted to playgroup because no detailed information on the number of sessions per week and age at start and finish of other informal care, such as friends or grandparents.

Children reported as attending special education settings, a crèche, or other settings were removed from the sample due to low numbers. The third exposure was age at start: “4-5”, “3-3.99”, or “0-2.99” years old when started. The fourth exposure was intensity: “1 day/week” if attending 1-2 sessions, “2 days/week” for 3-4 sessions, “3 days/week” for 5-6 sessions, or “4-5 days/week” for 7-10 sessions. When the current placement started after five years of age, or a mother reported that their child did not attend childcare, or data were missing, we used the information from the most recent previous placement (with complete data) to derive exposures two to four.

#### Potential confounders

In addition to sex (female vs male), Father’s occupational class and mother’s age of leaving full time education were considered as proxies of socio-economic position (SEP). Father’s occupation, measured when the participant was 10 years old, was classified according to the Registrar General’s Social Class (I professional, II managerial and technical, IIIN skilled non-manual, IIIM skilled manual, IV partly-skilled, and V unskilled).(20) Mother’s age at leaving full time education was ascertained at the birth sweep, and was categorised as ≥ 17 years old, 16 years old, 15 years old, or ≤ 14 years old.

### Statistical analyses

Descriptive statistics were produced. Further, in order to understand the interrelatedness of childcare type, age at start, and intensity, these three categorical variables were tabulated against each other. Chi-squared tests were used to quantify the strength of association between any given two variables.

#### Trajectory modelling

BMI trajectories were modelled in a multilevel general linear regression framework (measurement occasion at level one and individuals at level two),(21,22) incorporating systematic differences in the sample-average trajectory according to the childcare exposures and adjustment for potential confounders.

The time scale was decimal years of age modelled as a linear spline with a single knot at 26 years of age, thereby producing three easily interpretable parameters: 1) BMI (kg/m^2^) at age 10 years, 2) BMI change (kg/m^2^/year) between ages 10-26 years, and 3) BMI change (kg/m^2^/year) between ages 26-42 years. The constant and two spline terms were allowed to have random effects at level two, with an unstructured variance-covariance matrix. Further, the level one variance (i.e., error) was allowed to differ according to whether the data were measured or self-reported. Placing the knot at the other logical point (i.e., age 30 years) or using more complex functions, including fractional polynomials, did not result in noticeably better fitting models.

#### Individual childcare exposures

In the first set of models, each of the four childcare exposures was considered separately and was included as a main effect and as interactions with the two spline terms, thereby producing estimates capturing the association of the exposure with BMI at age 10 years, BMI change between ages 10-26 years, and BMI change between ages 26-42 years. Sex, father’s occupational class, and mother’s age at leaving full time education were included in the same way to provide robust adjustment for these potential confounders. For parsimony, the two SEP variables were converted and entered into models as ridit scores; associated estimates capture the difference in BMI between the lowest and highest SEP. The models were also used to estimate associations of the exposures with BMI at ages 26 and 42 years. No strong evidence of effect modification by SEP was found in stratified models or in models incorporating exposure-by-SEP interaction terms.

#### Different childcare age at start and intensity combinations

Because childcare type, age at start, and intensity were related to each other, it was not prudent to fit a single model that included mutual adjustment for these three variables. Instead, in the second set of models, the relationships of different age at start and intensity of childcare combinations with BMI trajectories were investigated in all children who were reported to attend childcare. The childcare intensity variable was collapsed and combined with the age at start of childcare variable to create a new exposure with six responses: 1) 4-5 years old when started and attended 1-2 days/week, 2) 4-5 years old when started and attended 3-5 days/week, 3) 3-3.99 years old when started and attended 1-2 days/week, 4) 3-3.99 years old when started and attended 3-5 days/week, 5) 0-2.99 years old when started and attended 1-2 days/week, and 6) 0-2.99 years old when started and attended 3-5 days/week. This exposure was entered into a single model in the same way that each individual exposure was investigated in the first set of models, with the same set of adjustments for potential confounders. Again, no strong evidence of effect modification, this time by childcare type (i.e., informal vs formal), was observed in exploratory analyses.

All procedures were performed in Stata 15 (StataCorp LP, College Station, TX, USA). The command runmlwin was used for the multilevel models.(23)

## Results

Approximately 85% of the sample reported using childcare, with a preference for informal over formal childcare (62 vs 38%) (Table 1). Among those who attended any childcare, the majority (61%) started between 3-4 years of age and the largest proportion (36%) attended for just one day a week. As shown in Table 2, however, age at start and intensity differed according to type of childcare, with children in formal childcare tending to start at a later age and attend more times per week than children who attended informal childcare. Conversely, those children who started at a later age tended to go more times per week.

**Table 1.** Description of sample of 8234 participants

|  |  |  |
| --- | --- | --- |
| Childcare |  |  |
| No | N (%) | 1265 (15.4) |
| Yes | N (%) | 6969 (84.6) |
| Type |  |  |
| Formal | N (%) | 2634 (37.8) |
| Informal | N (%) | 4335 (62.2) |
| Age at start |  |  |
| 4-5 years old when started | N (%) | 1539 (22.1) |
| 3-3.99 years old when started | N (%) | 4216 (60.5) |
| 0-2.99 years old when started | N (%) | 1214 (17.4) |
| Intensity |  |  |
| 1 day/week | N (%) | 2510 (36.0) |
| 2 days/week | N (%) | 1601 (23.0) |
| 3 days/week | N (%) | 1995 (28.6) |
| 4-5 days/week | N (%) | 863 (12.4) |
| Sex |  |  |
| Male | N (%) | 4188 (50.9) |
| Female | N (%) | 4046 (49.1) |
| Ethnicity |  |  |
| White British | N (%) | 7930 (96.3) |
| Other | N (%) | 255 (3.1) |
| Missing | N (%) | 49 (0.6) |
| Father's occupational class at age 10 years |  |  |
| I (Professional) | N (%) | 451 (5.5) |
| II (Managerial and technical) | N (%) | 2028 (24.6) |
| IIIN (Skilled non-manual) | N (%) | 942 (11.4) |
| IIIM (Skilled manual) | N (%) | 3307 (40.2) |
| IV (Partly-skilled) | N (%) | 1028 (12.5) |
| V (Unskilled) | N (%) | 478 (5.8) |
| Age mother left full time education |  |  |
| ≥ 17 years old | N (%) | 1612 (19.6) |
| 16 years old | N (%) | 1490 (18.1) |
| 15 years old | N (%) | 4698 (57.1) |
| ≤ 14 years old | N (%) | 434 (5.3) |
| BMI (kg/m<sup>2</sup>) |  |  |
| Age 10 years (7478 observations) | Mean (SD) | 16.9 (2.1) |
| Age 16 years (4654 observations) | Mean (SD) | 21.1 (3.0) |
| Age 26 years (4138 observations) | Mean (SD) | 23.6 (3.7) |
| Age 30 years (6054 observations) | Mean (SD) | 24.8 (4.2) |
| Age 34 years (5330 observations) | Mean (SD) | 25.8 (4.6) |
| Age 42 years (4909 observations) | Mean (SD) | 26.8 (5.1) |

**Table 2.** Tabulations between childcare type, age at start, and intensity among 6969 participants

|  |  | Type |  | Intensity |  |  |  |
| --- | --- | --- | --- | --- | --- | --- | --- |
|  |  | Formal<br>N = 2634 | Informal<br>N = 4335 | 1 day/week<br>N = 2510 | 2 days/week<br>N = 1601 | 3 days/week<br>N = 1995 | 4-5 days/week<br>N = 863 |
| Age at start |  |  |  |  |  |  |  |
| 4-5 years old when started | N (%) | 952 (36.1) | 587 (13.5) | 414 (16.5) | 179 (11.2) | 615 (30.8) | 331 (38.4) |
| 3-3.99 years old when started | N (%) | 1360 (51.6) | 2856 (65.9) | 1687 (67.2) | 1029 (64.3) | 1108 (55.5) | 392 (45.4) |
| 0-2.99 years old when started | N (%) | 322 (12.2) | 892 (20.6) | 409 (16.3) | 393 (24.6) | 272 (13.6) | 140 (16.2) |
| Intensity |  |  |  |  |  |  |  |
| 1 day/week | N (%) | 178 (6.8) | 2332 (53.8) |  |  |  |  |
| 2 days/week | N (%) | 208 (7.9) | 1393 (32.1) |  |  |  |  |
| 3 days/week | N (%) | 1432 (54.4) | 563 (13.0) |  |  |  |  |
| 4-5 days/week | N (%) | 816 (31.0) | 47 (1.1) |  |  |  |  |

Chi-squared tests for each of the three tabulations had $p < 0.001$

### Individual childcare exposures

Table 3 shows the estimated associations of each childcare exposure with BMI trajectories from 10-42 years of age. Children who attended childcare did not have higher BMI at age 10 years than those who did not attend childcare (β 0.039 kg/m^2^; 95% CI -0.103, 0.180), and there was no evidence that this association changed over follow-up. When childcare was separated into “formal” and “informal” we similarly found no evidence that these groups had different BMI trajectories compared to children who did not attend childcare. Results for the age at start exposure did show that the BMI of participants who started childcare at 0-2.99 years of age was 0.266 kg/m^2^ (95% CI 0.095, 0.437) higher at 10 years of age than participants who started childcare at 4-5 years of age. But they subsequently gained less BMI between ages 10-26 years (β -0.013 kg/m^2^/year; 95% CI -0.032, 0.006) such that effect sizes were attenuated and null at ages 26 and 42 years. For the intensity exposure, however, attending childcare 4-5 days a week (compared to 1 day a week) resulted in higher BMI at age 10 years (β 0.170 kg/m^2^; 95% CI -0.006, 0.058) and this effect persisted to age 26 years (β 0.349 kg/m^2^; 95% CI 0.024, 0.673) and age 42 years (β 0.380 kg/m^2^; 95% CI -0.066, 0.826).

**Table 3.**
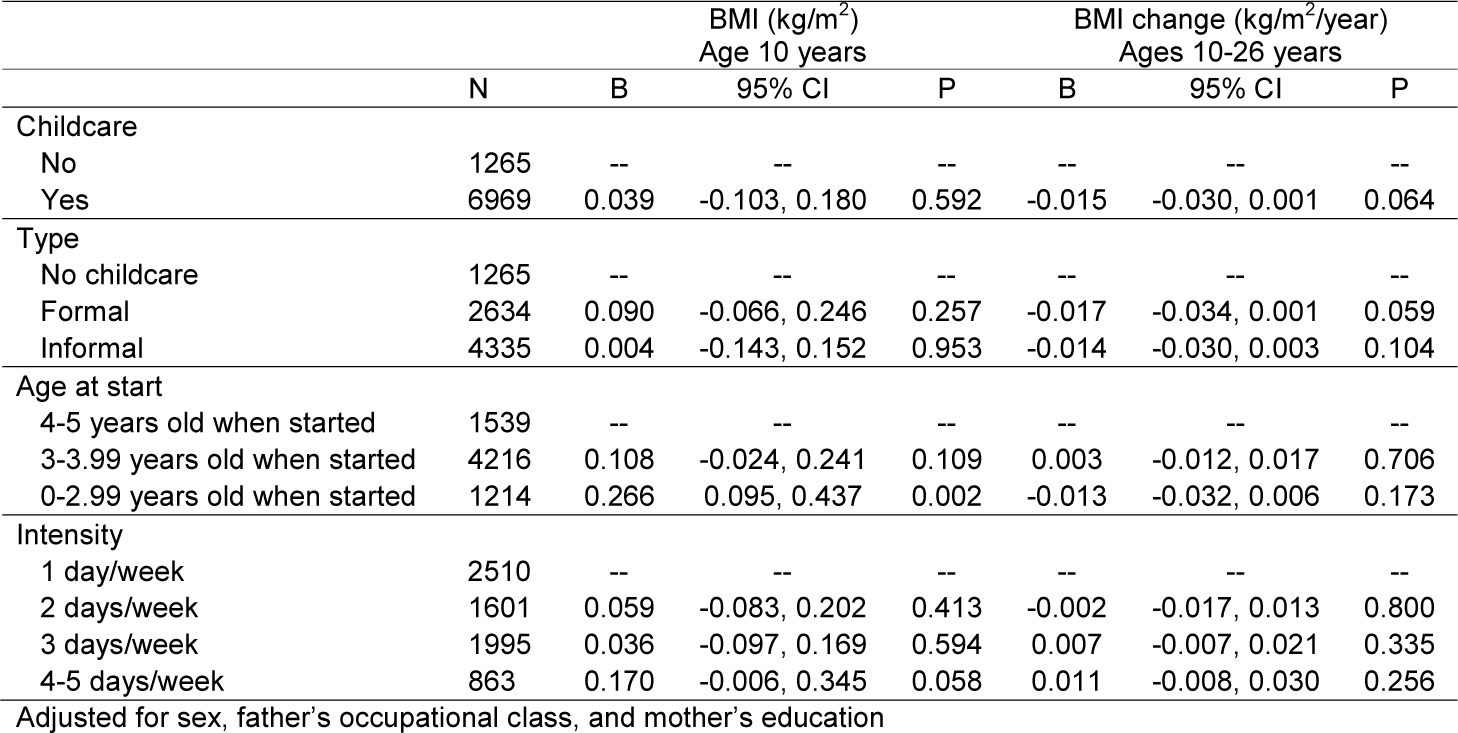
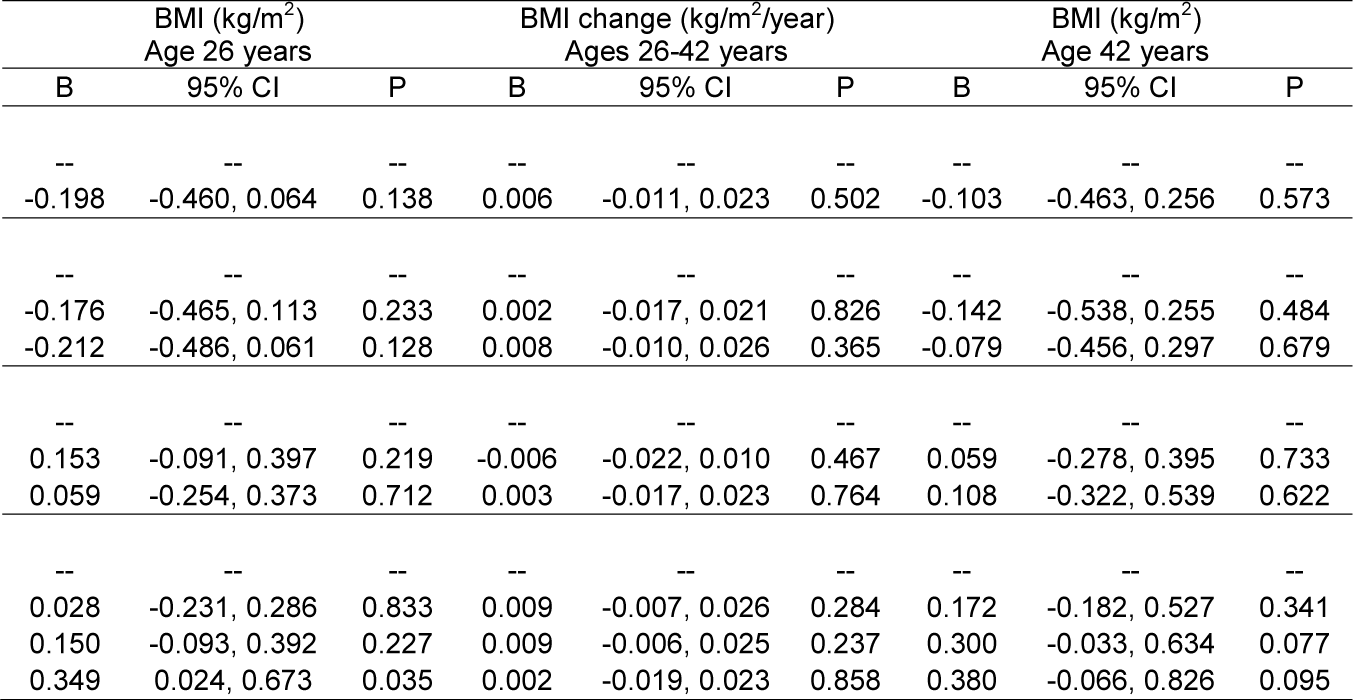
Associations of childcare type, age at start, and intensity with BMI trajectories from ages 10-42 years, among 8234 participants, estimated from four separate multilevel linear spline models

|  | N | BMI (kg/m<sup>2</sup>)<br>Age 10 years |  |  | BMI change (kg/m<sup>2</sup>/year)<br>Ages 10-26 years |  |  |
| --- | --- | --- | --- | --- | --- | --- | --- |
|  |  | B | 95% CI | P | B | 95% CI | P |
| Childcare |  |  |  |  |  |  |  |
| No | 1265 | -- | -- | -- | -- | -- | -- |
| Yes | 6969 | 0.039 | -0.103, 0.180 | 0.592 | -0.015 | -0.030, 0.001 | 0.064 |
| Type |  |  |  |  |  |  |  |
| No childcare | 1265 | -- | -- | -- | -- | -- | -- |
| Formal | 2634 | 0.090 | -0.066, 0.246 | 0.257 | -0.017 | -0.034, 0.001 | 0.059 |
| Informal | 4335 | 0.004 | -0.143, 0.152 | 0.953 | -0.014 | -0.030, 0.003 | 0.104 |
| Age at start |  |  |  |  |  |  |  |
| 4-5 years old when started | 1539 | -- | -- | -- | -- | -- | -- |
| 3-3.99 years old when started | 4216 | 0.108 | -0.024, 0.241 | 0.109 | 0.003 | -0.012, 0.017 | 0.706 |
| 0-2.99 years old when started | 1214 | 0.266 | 0.095, 0.437 | 0.002 | -0.013 | -0.032, 0.006 | 0.173 |
| Intensity |  |  |  |  |  |  |  |
| 1 day/week | 2510 | -- | -- | -- | -- | -- | -- |
| 2 days/week | 1601 | 0.059 | -0.083, 0.202 | 0.413 | -0.002 | -0.017, 0.013 | 0.800 |
| 3 days/week | 1995 | 0.036 | -0.097, 0.169 | 0.594 | 0.007 | -0.007, 0.021 | 0.335 |
| 4-5 days/week | 863 | 0.170 | -0.006, 0.345 | 0.058 | 0.011 | -0.008, 0.030 | 0.256 |

Table 3. Continued
| BMI (kg/m<sup>2</sup>)<br>Age 26 years |  |  | BMI change (kg/m<sup>2</sup>/year)<br>Ages 26-42 years |  |  | BMI (kg/m<sup>2</sup>)<br>Age 42 years |  |  |
| --- | --- | --- | --- | --- | --- | --- | --- | --- |
| B | 95% CI | P | B | 95% CI | P | B | 95% CI | P |
| --- | --- | --- | --- | --- | --- | --- | --- | --- |
| -0.198 | -0.460, 0.064 | 0.138 | 0.006 | -0.011, 0.023 | 0.502 | -0.103 | -0.463, 0.256 | 0.573 |
| --- | --- | --- | --- | --- | --- | --- | --- | --- |
| -0.176 | -0.465, 0.113 | 0.233 | 0.002 | -0.017, 0.021 | 0.826 | -0.142 | -0.538, 0.255 | 0.484 |
| -0.212 | -0.486, 0.061 | 0.128 | 0.008 | -0.010, 0.026 | 0.365 | -0.079 | -0.456, 0.297 | 0.679 |
| --- | --- | --- | --- | --- | --- | --- | --- | --- |
| 0.153 | -0.091, 0.397 | 0.219 | -0.006 | -0.022, 0.010 | 0.467 | 0.059 | -0.278, 0.395 | 0.733 |
| 0.059 | -0.254, 0.373 | 0.712 | 0.003 | -0.017, 0.023 | 0.764 | 0.108 | -0.322, 0.539 | 0.622 |
| --- | --- | --- | --- | --- | --- | --- | --- | --- |
| 0.028 | -0.231, 0.286 | 0.833 | 0.009 | -0.007, 0.026 | 0.284 | 0.172 | -0.182, 0.527 | 0.341 |
| 0.150 | -0.093, 0.392 | 0.227 | 0.009 | -0.006, 0.025 | 0.237 | 0.300 | -0.033, 0.634 | 0.077 |
| 0.349 | 0.024, 0.673 | 0.035 | 0.002 | -0.019, 0.023 | 0.858 | 0.380 | -0.066, 0.826 | 0.095 |

### Different age at start and intensity of childcare combinations

Table 4 shows the estimated associations of different age at start and intensity of childcare combinations with BMI trajectories. At 10 years of age, there was a clear dose-response relationship between greater exposure to childcare and increased BMI. For example, among participants who attended childcare 1-2 days a week, those who started when 3-3.99 years old had a 0.197 kg/m^2^ (95%CI -0.004, 0.399) higher BMI than those who started when 4-5 years old, and those who started when 0-2.99 years old had a 0.289 kg/m^2^ (95%CI 0.049, 0.529) higher BMI. Similarly, when holding age at start of childcare constant, estimated effects sizes were always larger for participants who attended childcare 3-5 days a week compared to those who attended 1-2 days a week. These results clearly demonstrate that age at start and intensity of childcare had additive effects on BMI in late childhood. Subsequently, at ages 26 and 42 years, the effect sizes were not as well ranked according to the level of exposure to childcare. Nonetheless, even at 42 years of age, individuals with the highest level of exposure (i.e., 0-2.99 years old when started and attended 3-5 days/week) had a 1.356 kg/m^2^ (95% CI 0.637, 2.075) higher BMI than individuals with the lowest level of exposure (i.e., 4-5 years old when started and attended 1-2 days/week).

**Table 4.**
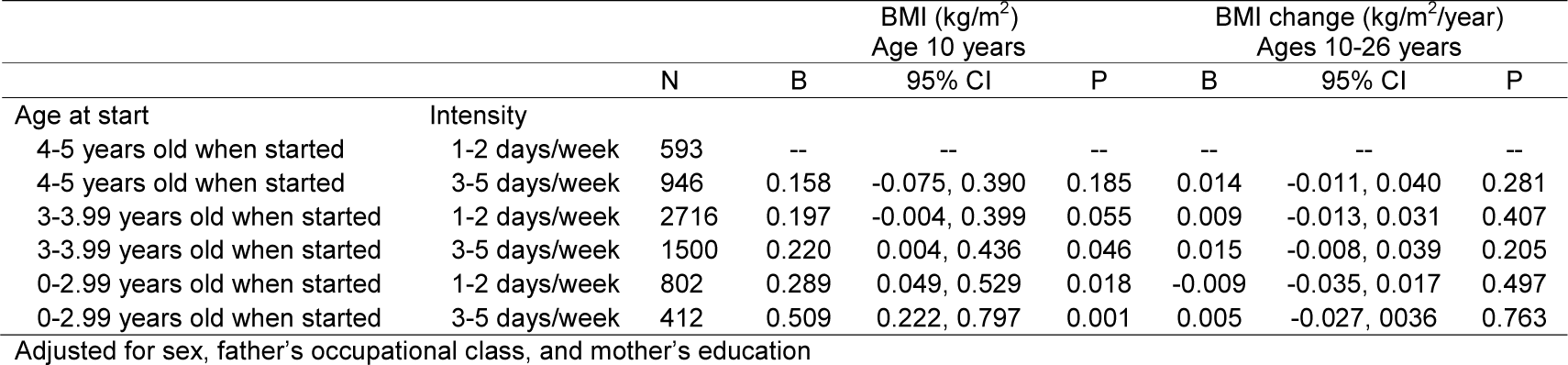
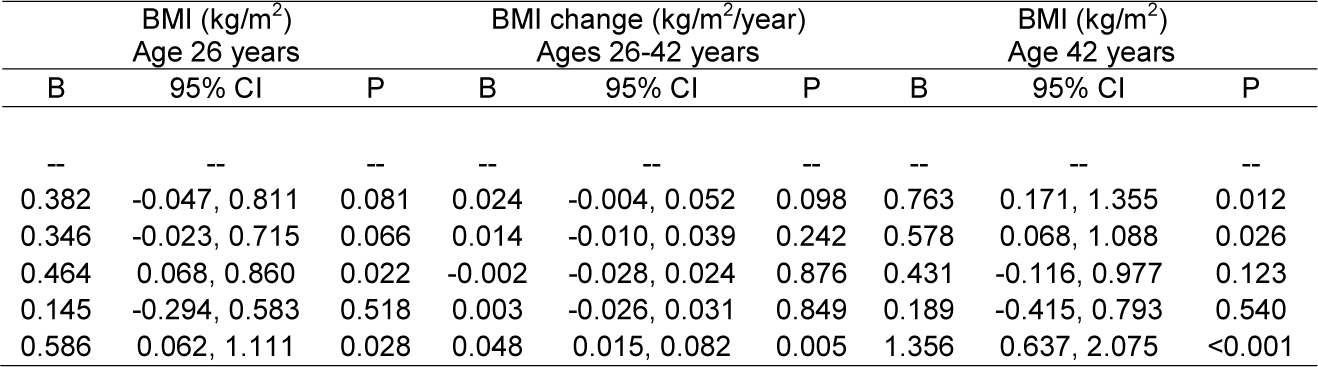
Associations of childcare age at start and intensity with BMI trajectories from ages 10-42 years, among 6969 participants, estimated from one single multilevel linear spline model

## Discussion

Using data from a large UK birth cohort study, our results demonstrate a clear dose-response relationship between starting childcare earlier and attending more frequently with higher BMI at age 10 years, in line with the findings of the Black et al systematic review.(18) While this finding is important in and of itself, the real gap in the literature is that, after childhood, we know almost nothing about how exposure to childcare might be related to BMI trajectories. The key finding of the present paper is that greater exposure to childcare resulted in higher estimated BMI up until age 42 years, perhaps with intensity being more deleterious than age at start of childcare attendance. As such, our paper provides the first evidence that targeting childcare policies and practices might ultimately benefit adulthood health and wellbeing.

Individuals with the highest level of childcare exposure were estimated to have between a 0.5-1.5 kg/m^2^ higher BMI than individuals with the lowest level of exposure across the studied age range. This effect size corresponds to approximately 0.1-0.3 standard deviations (SDs), which is not negligible given that a BMI reduction of more than 0.20-0.25 SDs has been proposed to be clinically important.(24)

Previous research in another of the UK nationally-representative studies, the 2000 Millennium Cohort Study, reported that children in informal childcare were more likely to be overweight at age three years than those cared for by a parent, but this may have been driven by 75% of informal care in that sample being primarily by grandparents.(25) Other studies have also reported higher BMI in children cared for by grandparents.(26,27) Unfortunately, data on care by grandparents was not collected in the 1970 British Cohort Study. Mothers whose primary source of childcare was grandparents would have either selected “other” or chosen one of the other options (e.g., playgroup), and this might explain why we found no association between childcare type and BMI trajectories. The Millennium Cohort Study paper also found that the relationship of informal childcare with greater overweight risk was strongest among more advantaged groups (e.g., managerial or professional background).(25) This might be because, with higher SEP, childcare increasingly represents a less healthy environment than that found at home. We found no formal evidence of effect modification by father’s occupational class and mother’s age of leaving full time education, but this may reflect the lower power in these sub-group analyses. While the 1970 British Cohort Study offers the opportunity to model childhood to adulthood BMI trajectories, incorporating effect modification into these longitudinal models requires three-way interactions and thus a very large sample size. The Millennium Cohort Study now has data to age 14 years and would allow detailed investigation with many different outcomes in a contemporaneous sample.

There are only two studies that we are aware of that have investigated the association of childcare with BMI in adulthood. In a sample of 783 women attending Cracow or Opole universities in Poland in 2005, Wronka and Pawlinnska-Chmara found no association of childcare type with BMI at 20-24 years of age.(28) In another of the UK nationally-representative studies, the 1958 National Child Development Study, Batty et al found that the BMI at age 44/45 years of individuals who attended nursery was only 0.01 (-0.05, 0.07) SDs higher compared to individuals who did not attend nursery.(29) We similarly found no evidence of an association between childcare attendance or type with BMI at ages 42 years but did find evidence for childcare intensity. Batty et al also reported no association of nursery attendance with a range of cardio-metabolic disease risk factor (e.g., blood pressure and cholesterol) and mortality. Further work, however, needs to investigate the relationships of age at start and intensity of childcare with these and other outcomes.

The pathways through which childcare experiences may affect the risk of obesity are poorly understood.(26,30) Different types and characteristics of childcare providers may have different influences on the development of obesity-related risk factors, such as physical activity, sedentary behaviour, sleep, diet, and stress.(31-33) For example, a recent systematic review reported that some childcare staff behaviours (e.g. providing portable play equipment and positively prompting children to be active) were associated with increased physical activity in children in cross-sectional studies.(33) However, this was not seen across all studies. Given the existing evidence of associations between physical activity, sedentary behaviour, sleep, diet, and stress with increased adiposity in early childhood,(30,34) it has been hypothesised that these risk factors are possible pathways through which the childcare experience may influence the development of obesity.(35) However, the scarcity of published studies investigating the longitudinal associations of childcare during the early years and subsequent risk factors such as physical activity and sedentary behaviour limit the ability to confirm this hypothesis.

The main strengths of the present article are 1) the thorough analysis of longitudinal BMI data collected on a large cohort over three decades of follow-up and 2) the ability to investigate multiple aspects of childcare (i.e., attendance, type, age at start, and intensity). In terms of limitations, 1) childcare in the 1970’s may be very different from today, although analyses in more recent cohorts have also demonstrated deleterious consequences for short-term obesity risk,(25) thereby suggesting that our results may be relevant for contemporary childcare settings, 2) we were unable to investigate the potential effects of concurrent attendance to multiple childcare settings because data on age at start and intensity of attendance were only available for the main childcare setting attended, 3) BMI is only a limited indicator of adiposity and the reported associations may also reflect greater fat-free mass,(9,36,37) 4) despite adjustment for father’s occupational class and mother’s age of leaving full time education, we cannot rule out the possibility of residual confounding by SEP, and 5) we cannot assess or rule out reverse causality because BMI was not assessed before age 10 years in this cohort.

In conclusion, our results provide evidence that starting childcare at an earlier age and attending more frequently have additive effects on increasing BMI which persist from late-childhood to mid-adulthood. Use of childcare is both necessary and highly prevalent.(14-16) As such, strategic research is needed to understand what policies and practices regarding physical activity, sedentary behaviour, and diet exist and are implemented in these settings, and how these could be changed so that childcare plays a key role in the establishment of healthy behaviours and weight across the life course, instead of being associated with a more adverse BMI trajectory.

## Data Availability

Source data are available from the UK Data Archive

https://www.data-archive.ac.uk/

## Acknowledgments

WJ is supported by the UK Medical Research Council (New Investigator Research Grant: MR/P023347/1) and acknowledges support from the National Institute for Health Research (NIHR) Leicester Biomedical Research Centre, which is a partnership between University Hospitals of Leicester NHS Trust, Loughborough University, and the University of Leicester.

JA is funded by the Centre for Diet and Activity Research (CEDAR), a UKCRC Public Health Research Centre of Excellence which is funded by the British Heart Foundation, Cancer Research UK, Economic and Social Research Council, Medical Research Council, the National Institute for Health Research, and the Wellcome Trust (MRC administered grant MR/K023187/1).

WJ and SC conceptualized the study. WJ carried out the analyses and SC drafted the initial manuscript. SC, DB, SBN, JA, and WJ made substantial contributions to the interpretation of the data, revised the manuscript critically for important intellectual content, gave final approval of the version to be published, and agree to be accountable for all aspects of the work.

